# Identifying Signs and Symptoms of Urinary Tract Infection from Emergency Department Clinical Notes Using Large Language Models

**DOI:** 10.1101/2023.10.20.23297156

**Authors:** Mark Iscoe, Vimig Socrates, Aidan Gilson, Ling Chi, Huan Li, Thomas Huang, Thomas Kearns, Rachelle Perkins, Laura Khandjian, R. Andrew Taylor

## Abstract

**Objectives:** Symptom characterization is critical to urinary tract infection (UTI) diagnosis, but identification of symptoms from the electronic health record (EHR) is challenging, limiting large-scale research, public health surveillance, and EHR-based clinical decision support. We therefore developed and compared two natural language processing (NLP) models to identify UTI symptoms from unstructured emergency department (ED) notes.

**Methods:** The study population consisted of patients aged ≥ 18 who presented to the (ED) in a north-eastern United States health system between June 2013 and August 2021 and had a urinalysis performed. We annotated a random subset of 1,250 ED clinician notes from these visits for a list of 17 UTI symptoms. We then developed two task-specific large language models (LLMs) to perform the task of named entity recognition (NER): a convolutional neural network (CNN)-based model (SpaCy) and a transformer-based model designed to process longer documents (Longformer). Models were trained on 1,000 notes and tested on a holdout set of 250 notes. We compared model performance (precision, recall, F1 measure) at identifying the presence or absence of UTI symptoms at the note level.

**Results:** 8,135 entities were identified in 1,250 notes; 83.6% of notes included at least one entity. Overall F1 measure for note-level symptom identification weighted by entity frequency was 0.84 for the SpaCy model and 0.88 for the Longformer model. F1 measure for identifying presence or absence of any UTI symptom in a clinical note was 0.96 (232/250 correctly classified) for the SpaCy model and 0.98 (240/250 correctly classified) for the Longformer model.

**Conclusions:** The study demonstrated the utility of LLMs and transformer-based models in particular for extracting UTI symptoms from unstructured ED clinical notes; models were highly accurate for detecting the presence or absence of any UTI symptom on the note level, with variable performance for individual symptoms.

## Introduction

### Background

Antibiotic-resistant bacteria pose a significant public health threat, causing approximately 2 million illnesses and 35,000 deaths and incurring $20-35 billion in costs annually in the United States.[1] A primary contributor to this resistance is the inappropriate use of antibiotics, including over-prescribing and misaligned prescribing practices. This problem is particularly pronounced for the more than 2 million emergency department (ED) patients evaluated for urinary tract infections (UTIs) each year.[2] These patients often present with higher acuity, atypical symptoms, and worse baseline resistance patterns,[3] which increase the complexity of their cases.[4] As with other infections, UTIs with antibiotic resistance are more likely to progress to sepsis, result in prolonged hospital stays, and contribute to morbidity and mortality.[5, 6]

Compounding these issues is the fact that gold-standard diagnostic criteria from the CDC and the Infectious Diseases Society of America (ISDA) rely on microbial culture data, which is typically not available at the time of patient evaluation.[7] ED clinicians must therefore initiate antibiotics based on incomplete information, including patient symptoms, physical exam findings, and urinalysis results. Prior studies have indicated that improvements could be made in 30%–60% of cases regarding UTI diagnosis, treatment indications, agent choice, and antibiotic therapy duration in the ED setting.[4, 8] Many patients who are diagnosed with UTI and started on antibiotics in the ED ultimately have negative urine cultures, suggesting unnecessary treatment.[9–11] Other patients whose urine culture data is ultimately positive are treated with antibiotics despite the lack of signs or symptoms suggestive of infection[12]–such treatment of so-called “asymptomatic bacteriuria” poses its own risks, including antibiotic resistance, medication side effects, and increased length of stay.[13, 14] Once initiated, antibiotics are often continued inappropriately.[15] It is therefore desirable to improve the accuracy of UTI diagnosis in real time and to study UTI diagnosis and treatment at a large scale.

### Importance

Identification of patient signs (i.e., physical exam findings) and symptoms is essential for UTI diagnosis and the development of machine learning models for predicting and surveilling UTIs, as it allows for the differentiation of a UTI and asymptomatic bacteruria.[16] The documentation of UTI symptoms can be either structured (e.g., ICD-10 codes) or—as is often the case with signs and symptoms—unstructured (e.g., clinical notes), complicating symptom extraction. Natural language processing (NLP) has been employed to extract patient symptoms and diagnoses from clinical notes, including UTI symptoms from home care nursing notes.[17] However, well-validated NLP methods for UTI symptom extraction in the ED are lacking. Various NLP techniques, such as rule-based methods, machine learning, deep learning, and large language models (LLMs) have been applied in different healthcare contexts to extract valuable information from unstructured data.18 Techniques such as named entity recognition (NER) and text classification can be used to identify symptoms, extract relevant information, and classify clinical notes according to specific criteria. Despite these advancements, there remains a gap in understanding the characteristics of patients presenting with UTI symptoms in the ED and the potential of NLP techniques in this specific context.

### Goal of Investigation

The primary objective of this investigation is to assess the potential of NLP techniques in identifying UTI symptoms from unstructured clinical notes in the emergency department ED setting. In turn, we hope to facilitate research and public health surveillance by removing barriers to UTI sign and symptom identification from EHR databases and help pave the way for improved UTI diagnosis, treatment, and patient outcomes in the face of rising ED volumes, patient complexity, and antibiotic resistance.

## Methods

### Study Population and Setting

The study population consisted of adult patients (≥ 18 years of age) who presented to the emergency department (ED) between June 2013 and August 2021 and had a urinalysis sent during their visit. The study was conducted across ten sites within a regional healthcare network in the northeastern United States, covering a geographic area of approximately 650 square miles. The study followed the STROBE reporting guidelines for observational studies.[18] Our institutional review board approved this research and waived the need for informed consent (HIC# 1602017249).

### Data Collection and Processing

Patient demographic and clinical data were extracted from the system-wide electronic health record (Epic, Verona, WI) using a centralized data warehouse (Helix). The warehouse includes procedural instances (Epic procedural codes), prescription and medications, ICD-10 code diagnoses, laboratory records, and clinical notes.

### Defining UTI Signs and Symptoms

We identified a comprehensive set of likely and potential UTI signs and symptoms, identified through literature review,[19–21] society guidelines,[22, 23] and expert opinion (AT, MI). The following list of signs symptoms, presented in Table 1, served as the basis for our annotation process.

**Table 1:**
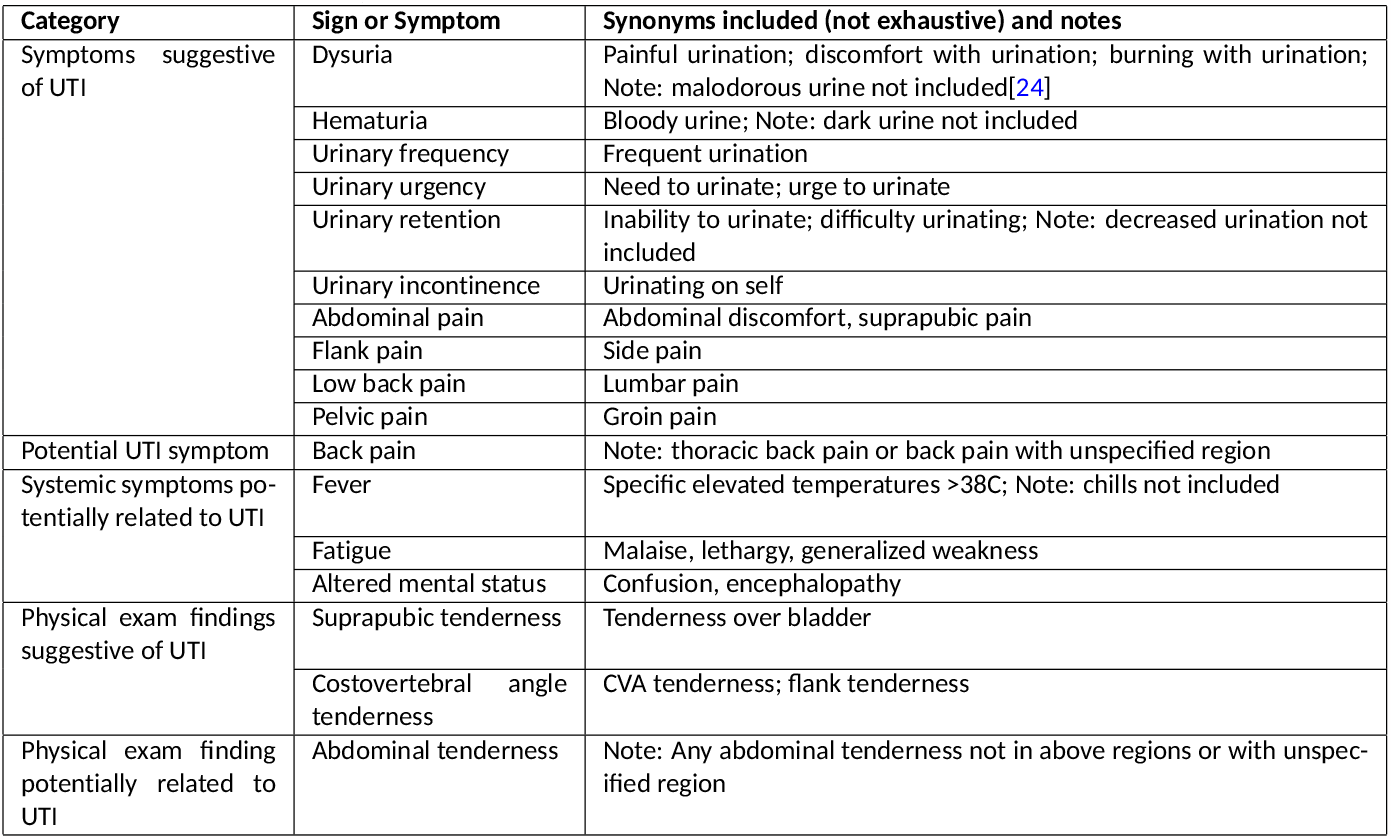
List of UTI signs and symptoms used for document annotation, along with a non-exhaustive list of synonyms and notes on sign or symptom definitions.

| Category | Sign or Symptom | Synonyms included (not exhaustive) and notes |
| --- | --- | --- |
| Symptoms suggestive of UTI | Dysuria | Painful urination; discomfort with urination; burning with urination; Note: malodorous urine not included[24] |
|  | Hematuria | Bloody urine; Note: dark urine not included |
|  | Urinary frequency | Frequent urination |
|  | Urinary urgency | Need to urinate; urge to urinate |
|  | Urinary retention | Inability to urinate; difficulty urinating; Note: decreased urination not included |
|  | Urinary incontinence | Urinating on self |
|  | Abdominal pain | Abdominal discomfort, suprapubic pain |
|  | Flank pain | Side pain |
|  | Low back pain | Lumbar pain |
|  | Pelvic pain | Groin pain |
| Potential UTI symptom | Back pain | Note: thoracic back pain or back pain with unspecified region |
| Systemic symptoms potentially related to UTI | Fever | Specific elevated temperatures >38C; Note: chills not included |
|  | Fatigue | Malaise, lethargy, generalized weakness |
|  | Altered mental status | Confusion, encephalopathy |
| Physical exam findings suggestive of UTI | Suprapubic tenderness | Tenderness over bladder |
|  | Costovertebral angle tenderness | CVA tenderness; flank tenderness |
| Physical exam finding potentially related to UTI | Abdominal tenderness | Note: Any abdominal tenderness not in above regions or with unspecified region |

### Document Annotation

We employed 17 labels for symptom and sign extraction: dysuria, hematuria, urinary frequency, urinary urgency, urinary incontinence, urinary retention, abdominal pain, flank pain, pelvic pain, low back pain, back pain, fever, fatigue, altered mental status, suprapubic tenderness, costovertebral angle tenderness (CVA), and abdominal tenderness (Figure **??**). The process began with the assembly of a diverse and representative set of ED provider notes from the above visits; notes were authored by an attending physician, an attending physician and resident physician jointly, an advanced practice provider (APP; a physician assistant, nurse practitioner, or advanced practice registered nurse) alone, or an APP and attending physician. Our note dataset was randomly selected and enriched for patients with UTI diagnoses and admitted patients so that it included, in roughly equal parts, patients who were admitted with a UTI diagnosis (see Table S1 for definitions), patients who were admitted without a UTI diagnosis, patients who were discharged with a UTI diagnosis, and patients who were discharged without a UTI diagnosis.

During an initial review of notes, we noticed that UTI signs or symptoms were localized to individual spans of text. Therefore, we framed the UTI symptom annotation task as a classic manual named entity recognition (NER) inside-outside-beginning (IOB) labeling task. However, given that the goal of our study was to train an NLP pipeline to identify patients with particular UTI signs or symptoms, we evaluated our annotation and model performance as a multilabel classification task, at the note level (i.e., our evaluation only considers the presence/absence of a particular symptom in the clinical note as a whole, not the particular text sequence that identifies the symptom). The annotation process began with the initial manual annotation of a set of 100 clinical notes by both MI and AT; MI and AT then adjudicated any conflicting annotations to create a “gold standard” annotation set through which to evaluate additional annotators. Following the initial manual annotation, a human-in-the-loop approach was employed to train the model on the remaining annotations. Our team of additional experienced annotators, comprising senior emergency medicine resident physicians, underwent thorough training on annotation guidelines developed by MI and AT including the notes and definitions in Table 1. Each annotator then annotated the gold-standard set of 100 provider notes adjudicated by MI and AT; their performance was assessed by measuring inter-annotator agreement using support-weighted average F1 measures (harmonic mean of precision and recall). This metric allowed us to evaluate the consistency between annotators, ensuring a reliable and standardized annotation process throughout the study. Annotators with an overall F1 measure of >0.8 across annotations were allowed to proceed to independently annotate notes after receiving feedback on their initial annotations. During the annotation process, we maintained open communication channels among annotators to facilitate discussion, address uncertainties, and resolve ambiguities.

### Annotation Environment

For annotation we employed Prodigy v1.11.7., a scriptable annotation tool designed to maximize efficiency, enabling data scientists to perform the annotation tasks themselves and facilitating rapid iterative development in natural language processing (NLP) projects. Prodigy uses transfer learning to develop models with fewer examples, streamlining data collection for NLP projects. Its active learning system focuses on ambiguous model examples to improve annotation efficiency.

### NLP Development

As discussed, we framed the identification of UTI symptoms in clinical text as an NER task and fine-tuned two state-of-the-art LLMs to identify named entities labeled in the text as UTI signs or symptoms. We focus on two task-specific LLMs in particular: a conventional convolutional neural network (CNN)-based SpaCy model, and Clinical Longformer,[25] a transformer-based model that can leverage the longer context windows common to clinical text and has been shown to outperform classic Bidirectional Encoder Representations from Transformers (BERT)-based models on a number of information extraction tasks.[25] Training these two methods allows for comparison between classic non-transformer based methods and the state-of-the-art in information extraction of long clinical texts. Transformer-based models have demonstrated exceptional performance in various NLP tasks by capturing complex contextual information from large-scale text corpora, but require a large number of annotations, while simpler methods such as SpaCy may perform better under label-constrained settings. The SpaCy pipeline is an ensemble of a bag-of-words-based and a CNN-based model. The Clinical Longformer was initialized from the general domain Longformer, trained on English Wikipedia, open-source books, and news articles. It was further pretrained on MIMIC-III clinical notes to tailor the model to clinical texts. To adapt both models for the specific task of UTI sign and symptom identification, we employed a fine-tuning process that leveraged our annotated dataset consisting of emergency department provider notes. During the fine-tuning phase, the pre-trained models were further trained on the task-specific dataset, allowing them to learn the nuances of clinical text and UTI-related symptoms. The SpaCy model is randomly initialized and trains both the bag-of-words model and the CNN model from scratch based on the provided dataset. The Clinical Longformer model instead fine-tunes a multi-class classification layer on top of the base model. In the NER setting, both the SpaCy and Longformer models select a class from each of the IOB tags, for each UTI symptom (e.g. I-FEVER, B-DYSURIA), as well as an outside tag indicating a word not part of any UTI symptom (“O”), for a total of 35 classes. The Longformer model was fine-tuned using a multi-class classification layer added on top of the base model, with each class representing one of the UTI symptoms with an IOB tag or an additional class for non-symptom text. We used a cross-entropy loss function for training, which calculates the difference between the model’s predicted probability distribution and the true label distribution. The training process involved several iterations over an 80% split of the dataset (1,000 of 1,250 notes), stratified by the occurrence of certain UTI sign or symptom labels, with a pre-defined batch size, and gradient descent optimization was utilized to minimize the loss function.[26] The fine-tuned models were then evaluated on a held-out test set of 250 notes (20%) to assess its generalization capability and performance in identifying UTI signs and symptoms in unseen clinical text. To evaluate model performance, we post-processed the output of the NER pipelines, which identified references to specific named entities (single words, abbreviations, or phrases), to determine the existence of UTI symptoms within a particular note. This entailed calculating a binary variable for the model-identified presence or absence of a named entity at the note (as opposed to entity) level; for example, notes that contained 1 model-identified mention of “dysuria” or 5 model-identified mentions of “dysuria” would both be categorized as mentioning “dysuria”, while a note with no model-identified mentions of “dysuria” would not. We then employed standard evaluation metrics of precision, recall, and F1 measure at the note level to quantitatively measure the model’s performance in extracting UTI symptoms from the provider notes.

## Results

We identified 695,062 ED encounters from January 1st 2013 to January 24th 2022; 95,970 were excluded because the patient was under 18 at the time of the encounter, leaving a total of 599,092 encounters involving 295,207 total patients. Encounter and patient characteristics for the 1,250 encounters randomly selected for provider note annotation are shown in Table 2.

**Table 2:**
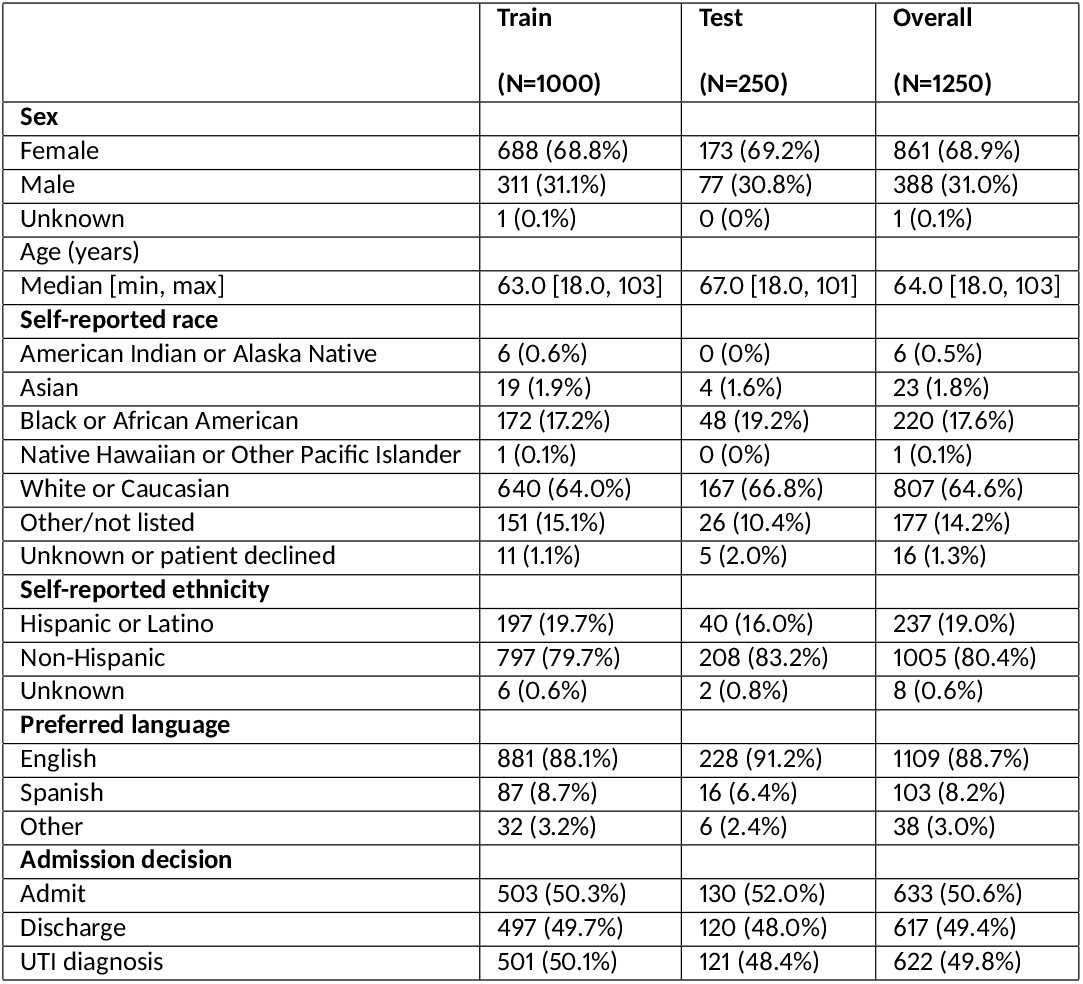
Baseline ED encounter and patient characteristics.

|  | Train<br>(N=1000) | Test<br>(N=250) | Overall<br>(N=1250) |
| --- | --- | --- | --- |
| **Sex** |  |  |  |
| Female | 688 (68.8%) | 173 (69.2%) | 861 (68.9%) |
| Male | 311 (31.1%) | 77 (30.8%) | 388 (31.0%) |
| Unknown | 1 (0.1%) | 0 (0%) | 1 (0.1%) |
| **Age (years)** |  |  |  |
| Median [min, max] | 63.0 [18.0, 103] | 67.0 [18.0, 101] | 64.0 [18.0, 103] |
| **Self-reported race** |  |  |  |
| American Indian or Alaska Native | 6 (0.6%) | 0 (0%) | 6 (0.5%) |
| Asian | 19 (1.9%) | 4 (1.6%) | 23 (1.8%) |
| Black or African American | 172 (17.2%) | 48 (19.2%) | 220 (17.6%) |
| Native Hawaiian or Other Pacific Islander | 1 (0.1%) | 0 (0%) | 1 (0.1%) |
| White or Caucasian | 640 (64.0%) | 167 (66.8%) | 807 (64.6%) |
| Other/not listed | 151 (15.1%) | 26 (10.4%) | 177 (14.2%) |
| Unknown or patient declined | 11 (1.1%) | 5 (2.0%) | 16 (1.3%) |
| **Self-reported ethnicity** |  |  |  |
| Hispanic or Latino | 197 (19.7%) | 40 (16.0%) | 237 (19.0%) |
| Non-Hispanic | 797 (79.7%) | 208 (83.2%) | 1005 (80.4%) |
| Unknown | 6 (0.6%) | 2 (0.8%) | 8 (0.6%) |
| **Preferred language** |  |  |  |
| English | 881 (88.1%) | 228 (91.2%) | 1109 (88.7%) |
| Spanish | 87 (8.7%) | 16 (6.4%) | 103 (8.2%) |
| Other | 32 (3.2%) | 6 (2.4%) | 38 (3.0%) |
| **Admission decision** |  |  |  |
| Admit | 503 (50.3%) | 130 (52.0%) | 633 (50.6%) |
| Discharge | 497 (49.7%) | 120 (48.0%) | 617 (49.4%) |
| UTI diagnosis | 501 (50.1%) | 121 (48.4%) | 622 (49.8%) |

### Annotation

MI, RT, and the three additional annotators each completed 250 annotations (in addition to the 100 gold standard annotations adjudicated by MI and RT and completed by each annotator for reliability testing), for a total of 1,250 annotations. The support-weighted macro-averaged F1 measures for the additional annotators compared to the gold standard was 0.82 (0.8-0.84). A total of 8,135 named entities were identified across the 1,250 notes; 83.6% of notes included at least one named entity. Details regarding frequency of named entities are shown in Table 3.

**Table 3:** UTI sign and symptom occurrence across 1,250 clinical notes, as identified through expert annotation.

| Category | Sign or Symptom | N (total occurrences) | Proportion of notes containing at least 1 occurrence of entity |
| --- | --- | --- | --- |
| Symptoms suggestive of UTI | Dysuria | 615 | 19% |
|  | Hematuria | 343 | 12% |
|  | Urinary frequency | 331 | 13% |
|  | Urinary urgency | 133 | 7% |
|  | Urinary retention | 171 | 6% |
|  | Urinary incontinence | 41 | 2% |
|  | Abdominal pain | 2226 | 38% |
|  | Flank pain | 600 | 13% |
|  | Low back pain | 116 | 5% |
|  | Pelvic pain | 165 | 6% |
| Potential UTI symptom | Back pain | 290 | 12% |
| Systemic symptoms potentially related to UTI | Fever | 862 | 19% |
|  | Fatigue | 642 | 22% |
|  | Altered mental status | 658 | 16% |
| Physical exam findings suggestive of UTI | Suprapubic tenderness | 104 | 6% |
|  | Costovertebral angle tenderness | 118 | 7% |
| Physical exam finding potentially related to UTI | Abdominal tenderness | 512 | 26% |
| Total (any entity) |  | 8135 | 84% |

### Model Performance

In the test set of 250 clinical notes, overall support-weighted model precision of the Longformer model was 0.86, recall was 0.92, and F1 measure was 0.88. Longformer model precision ranged from 0.58 for urinary retention to 1.00 for flank pain; recall ranged from 0.50 for urinary incontinence to 1.00 for low back pain and altered mental status; F1 measure ranged from 0.55 for urinary incontinence to 0.98 for flank pain. Overall support-weighted model precision of the SpaCy model was 0.89, recall was 0.83, and F1 measure was 0.84. SpaCy model precision ranged from 0 for urinary incontinence to 1.00 for urinary frequency, urinary urgency, altered mental status, and suprapubic tenderness; recall ranged from 0 for urinary incontinence to 0.98 for abdominal pain; F1 measure ranged from 0 for urinary incontinence to 0.96 for abdominal pain. Computing the macro-average precision, recall, and F1-scores across UTI symptoms, weighted by their prevalence in the dataset, allows us to account for label imbalance in our model evaluation. Details of model performance by named entity are shown in Table 4. Appendix Figure S1 shows confusion matrices for each model at the task of note-level identification of any UTI sign or symptom. Figure 2 provides a visualization of the trained Longformer model’s attention to various words and phrases in a deidentified clinical note.

**Table 4:**
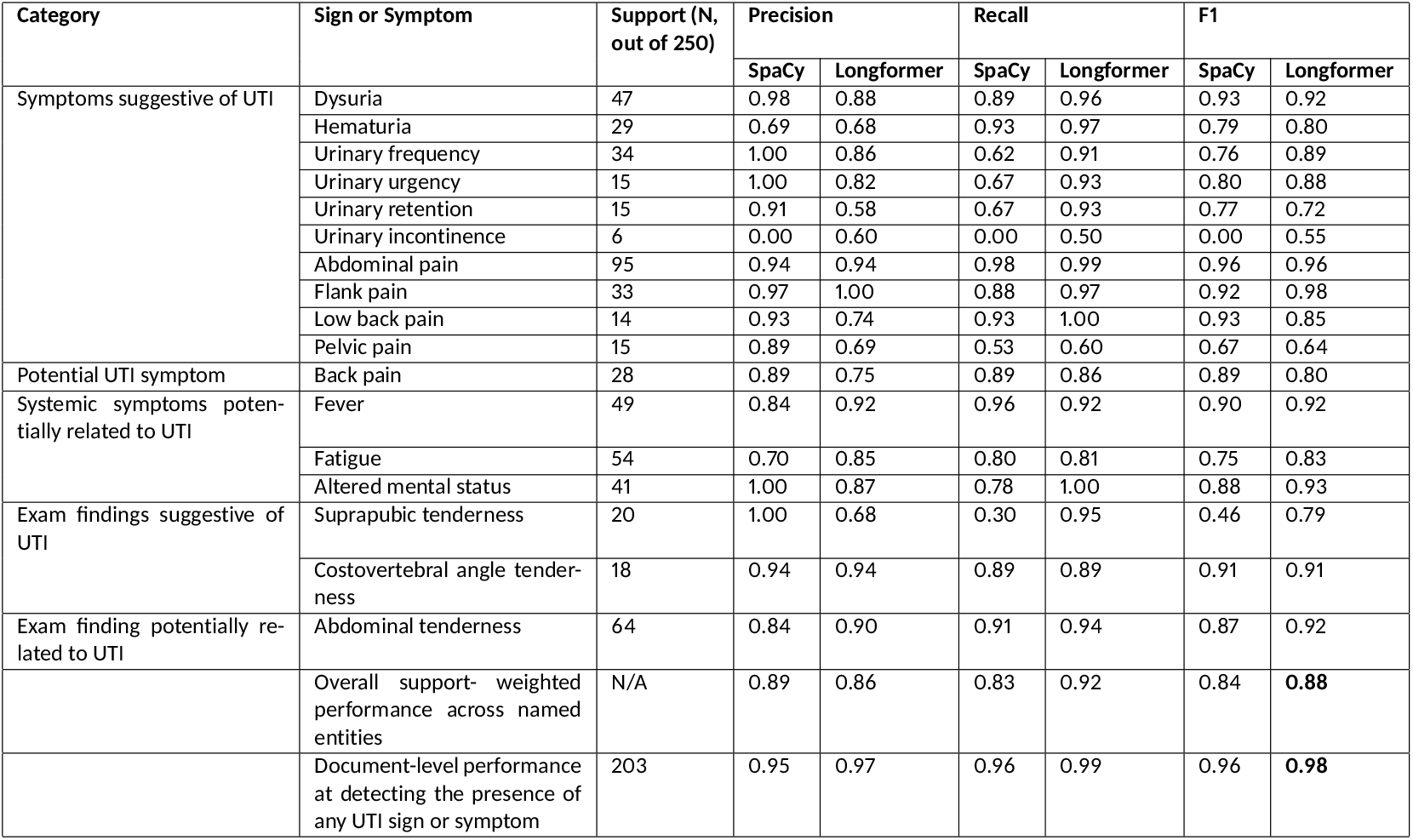
Longformer and SpaCy model performance by named entities across 250 notes in the test set. All values represent per-note entity recognition (i.e., model performance at identifying whether or not a note contains at least one occurrence of the named entity). For each named entity, the F1 value of the higher-performing model is underlined.

**Figure 1.**
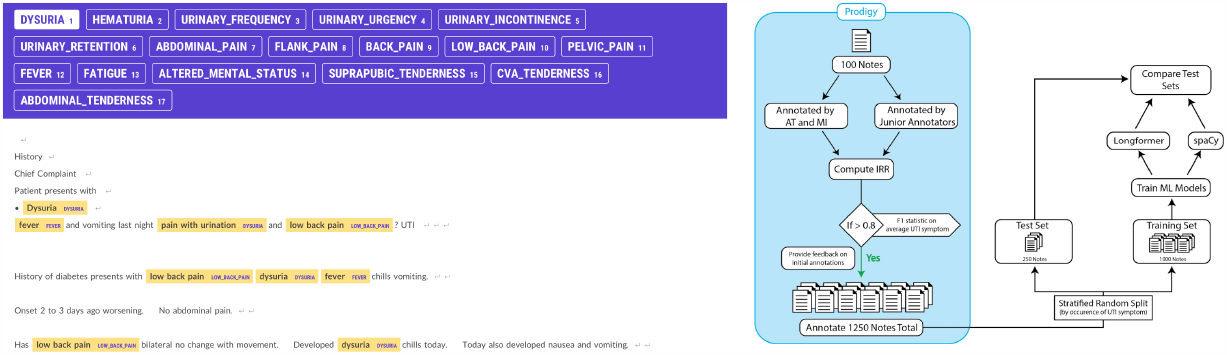
Image on the left shows a de-identified ED encounter note with several labeled entities highlighted in yellow in the Prodigy annotation interface. For each labeled entity, the original document text appears first, followed, in capital letters, by the entity label that has been applied. Note that “abdominal pain” has not been labeled as the presence of this symptom is negated in the note. Image on the right is a graphical abstract of the annotation, model training, and model evaluation process.

**Figure 2.**
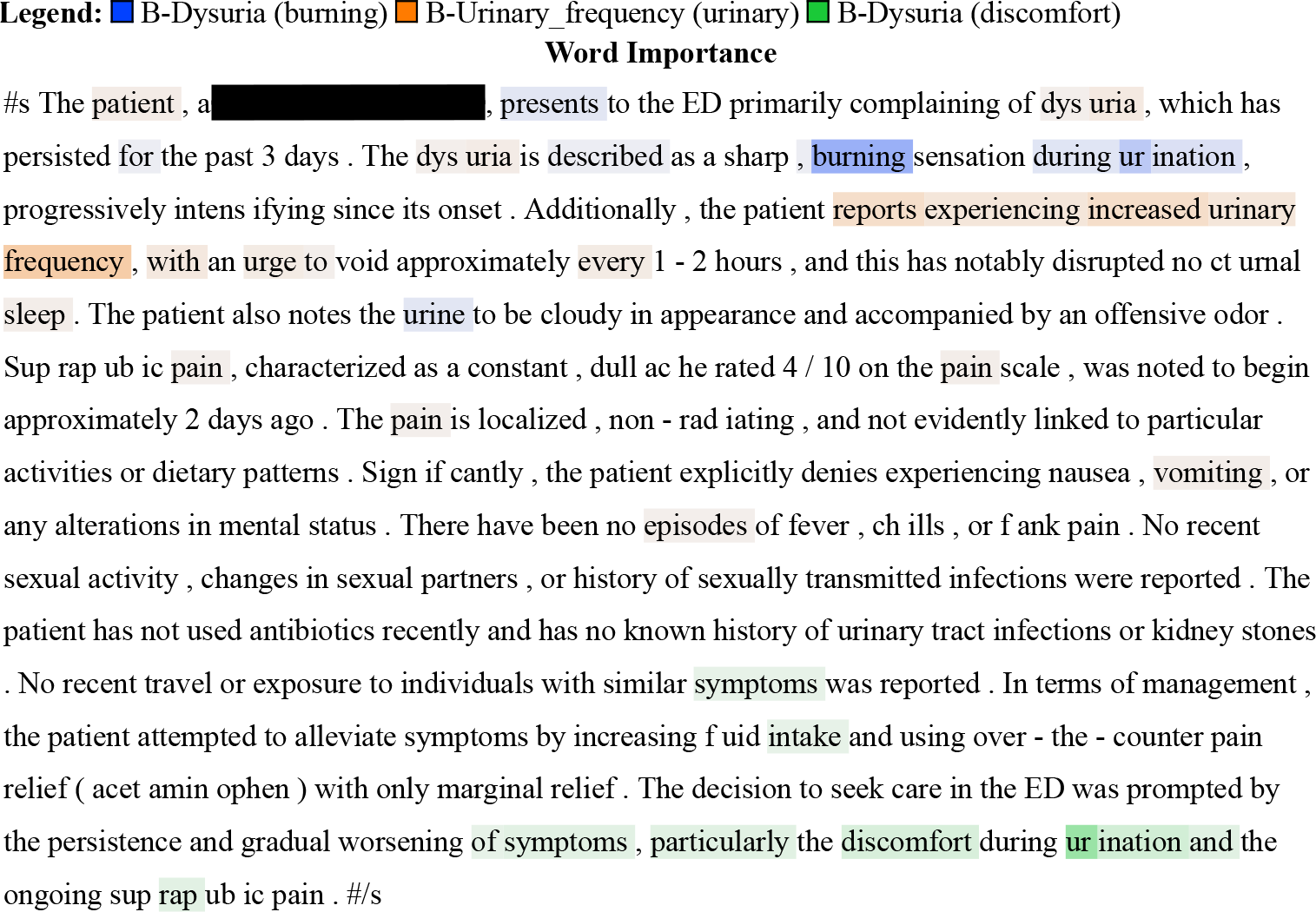
Visualization of relative importance of words and phrases in the trained Longformer model. Note that only three tokens–dysuria (burning), urinary frequency (urinary), and dysuria (discomfort), with parenthetical labels assigned by the model–have been included here for visual clarity. The legend identifies the color associated with each token; darker highlights connote greater relative importance.

## Discussion

Our study demonstrates that task-specific LLMs performing the NLP task of NER can effectively identify the presence or absence of UTI signs or symptoms in unstructured clinical notes from ED encounters. Because UTI diagnosis relies on signs and symptoms in addition to laboratory findings, such document-level classification is necessary to facilitate EHR-based UTI diagnosis on a large scale. These findings echo prior research describing NLP-based symptom information extraction from a variety of source documents.[27– 29] Model performance tended to be higher for entities with greater support (e.g., abdominal pain) and lower for those with lower support (e.g., urinary incontinence). Overall model performance at detecting the presence of any UTI sign or symptom was excellent for both models, with F1 measures of 0.96 and 0.98 for the standard-sequence CNN-based model (SpaCy) and long-sequence transformer-based model (Long-former), respectively, supporting the utility of NER for aiding in EHR-based UTI diagnosis. Consistent with prior research[25] showing improved performance of long-sequence transformer models (Longformer, Big-Bird) over standard-sequence models (RoBERTa, BERT) at clinical NER tasks, we found slightly better overall performance for our-long-sequence model. While the phrases referring to the named entities themselves tended to span several words at most, it is possible that the long-sequence model’s greater input length allowed for the incorporation of valuable contextual information such as the section of a note in which text appeared. A downside to long-sequence models is that they require greater memory and computational resources, potentially limiting real-time use.

### Challenges and Limitations

We faced several challenges that may be relevant to NLP-based symptom extraction tasks in other medical domains and contexts. First, because our dataset involved clinical notes with various formats and templates (related to clinical site, time of authorship, and author-specific practices), there were inconsistencies in the phrasing and verbiage surrounding documentation of signs or symptoms. Second, as in many medical NER tasks including a prior study using NLP to extract UTI-related information from home care nursing notes,[17, 30] we encountered frequent use of abbreviations, synonyms, and ambiguous terms (e.g., “AP” used to refer to “abdominal pain”–a named entity of interest–but also to “abdomen and pelvis” in computed tomography imaging, and “anteroposterior” in X-ray imaging, among numerous other concepts). Third, notes frequently included references to the absence of signs or symptoms, and we did not consider such references to negated entities to be instances of the named entities; we noted that the model performed poorly when words of negation (e.g., “denies” or “no”) were distant from the negated entity. The incorporation of a separate, dedicated “negation detection” model may improve model performance in NER tasks and will be explored in future work.[31] Finally, our dataset was highly imbalanced for the rarer entities. Because our primary objective was to classify documentation of any UTI sign or symptom–the information necessary to establish UTI diagnosis–rather than identification or enumeration of references to specific signs or symptoms, we elected against over-sampling low-frequency entities in model training; different sampling methods may be helpful in other clinical contexts. It should also be noted that EHR documentation of UTI signs and symptoms may not accurately represent patients’ experiences. Studies using patient surveys[32–34] and audio or video recordings[35, 36] of encounters have shown substantial discrepancies between patient-reported and physician-documented symptoms or exam findings. For example, a 2015 cross-sectional study of hospitalized patients with e. Coli bacteriuria found low correlation between UTI symptoms as reported in patient surveys and documented in emergency physicians’ (*κ*,0.09-0.5) and inpatient physicians’ (*κ*, 0.06-0.4) EHR notes.[37] While we did not quantify internal consistency in the notes we annotated, annotators anecdotally noted frequent internal discrepancies between symptom documentation in clinical notes with multiple authors (e.g., a resident physician and attending physician.)

### Future Directions

Future work should explore LLM performance in identifying UTI signs and symptoms from clinical notes from other care settings, such as inpatient medicine or ambulatory care, and in various patient groups. Similar methods could also be applied to signs and symptoms related to other illnesses and systems. Ultimately, this and similar models can facilitate accurate, large-scale extraction of UTI-related symptoms from unstructured clinical notes in the EHR, paving the way for research and public health surveillance examining the relationships among UTI signs and symptoms, urinalysis and urine culture results, UTI diagnoses, and patient outcomes. Better understanding of these relationships can help create systems that augment UTI diagnosis in real time, allowing for appropriately targeted treatment, improving patient outcomes, and limiting iatrogenic harm.

## Data Availability

All data produced in the present study are available upon reasonable request to the authors.

## Supplements

**Table S1:** List of ICD-10 codes for UTI diagnosis.

| IC-10 Code | Name |
| --- | --- |
| O03.38 | Urinary tract infection following incomplete spontaneous abortion |
| N10.XX | Acute pyelonephritis |
| N39.0 | Urinary tract infection, site not specified |
| A36.85 | Diphtheritic cystitis |
| A02.25 | Salmonella pyelonephritis |
| O86.20 | Urinary tract infection following delivery, unspecified |
| O23.40 | Unspecified infection of urinary tract in pregnancy, unspecified trimester |
| O08.83 | Urinary tract infection following an ectopic and molar pregnancy |
| N30.XX | Cystitis |
| N30.1 (ignored) | Interstitial Cystitis |

**Figure S1:**
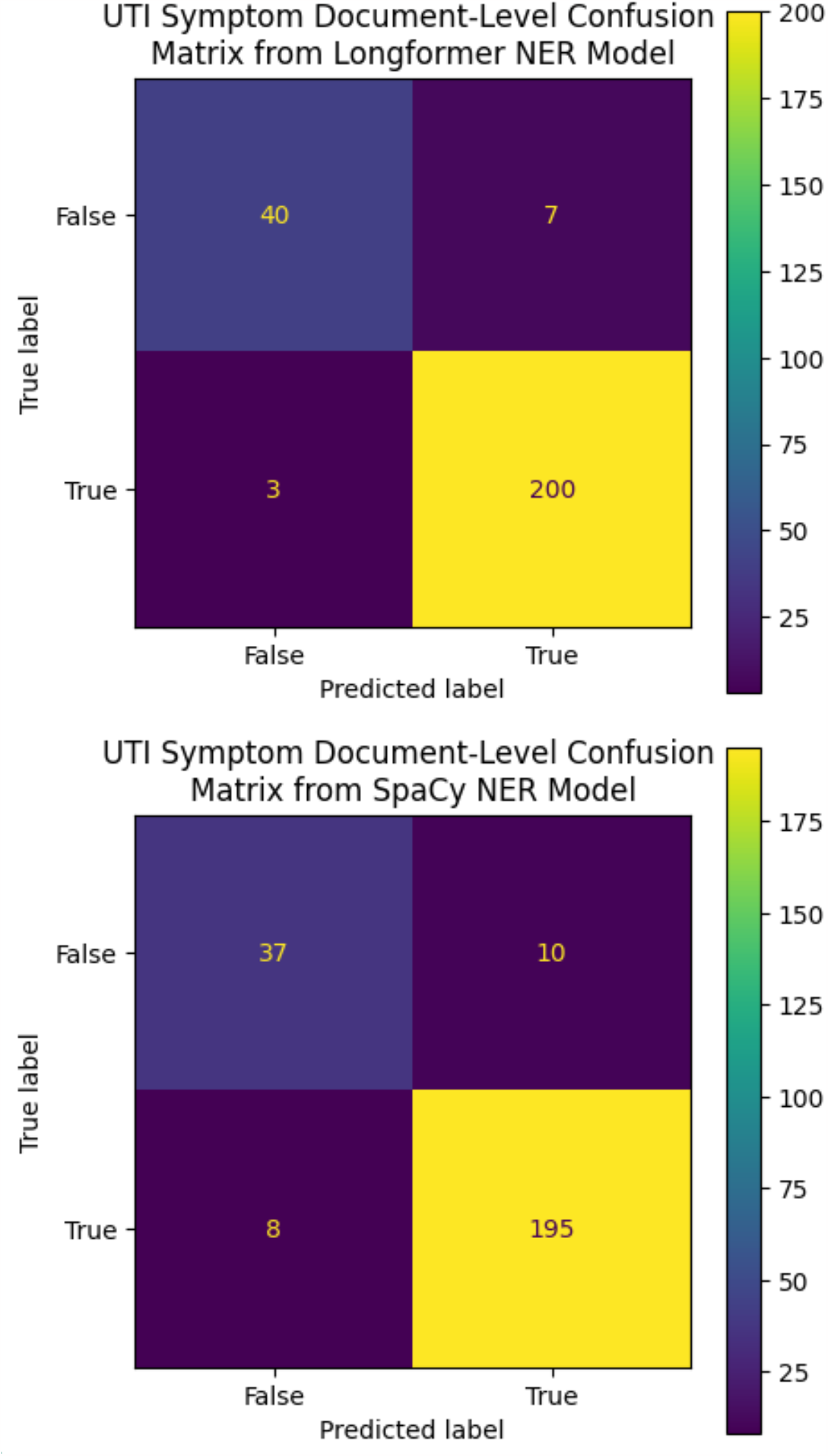
Confusion matrices showing (a) Longformer and (b) SpaCy model-predicted vs true labels for the presence or absence of any UTI sign or symptom at the note level.

